# Stool Testing for Tuberculosis Diagnosis in Pregnant Women and Under Five Children: A community based mixed method study from rural India

**DOI:** 10.64898/2026.09.03.26362132

**Authors:** Kajal Taluja, Shailja Shah, Arup Saha, Chandan Mahadevan, Navnit Nisha, Nazreen Nazir, Onkar Nath, Rakesh Kumar, Sweta Muni, Mithilesh Kumar, Ranjeet Kumar Sharma, Tushar Garg, Bal Krishna Mishra, Manish Kumar, Manish Bhardwaj, Miranda Brouwer

**Author notes:** Primary Corresponding Author: Chandan Mahadevan, Senior Project Manager, Innovators In Health (India), ‘Veer Sthal’ Ward No. 8, Keota Village, Dalsinghsarai, Samastipur District, Bihar, India-848114, Secondary Corresponding Author: Arup Saha, Research Manager, Innovators In Health (India), ‘Veer Sthal’ Ward No. 8, Keota Village, Dalsinghsarai, Samastipur District, Bihar, India-848114.

## Abstract

**Introduction:** Sputum sample is the standard for diagnosing pulmonary tuberculosis (TB) but many people, including pregnant women (PW) struggle to produce adequate amounts of sputum. *Mycobacterium tuberculosis (MTB)* persists in gastric fluid, thereby organisms in swallowed sputum may be detectable in stool.

**Objectives:** This study evaluates the utility of stool testing to improve TB diagnostic yield among pregnant women and under five children and explores factors affecting the feasibility of its implementation in programmatic settings.

**Methods:** We conducted a community based sequential mixed methods study between October 2023 to December 2024 in two districts of Bihar, India. PW and U5C were screened for TB at HWC-PHC. Those individuals, who were unable to expectorate sputum, were offered stool testing using Xpert MTB/RIF Ultra. We estimated the additional yield of TB by stool testing and explored the feasibility of its implementation across beneficiaries and implementors.

**Results:** A total of 21 PwTB were detected overall (10.3% (4/39) in PW and 80.9% (17/21) in U5C), through stool testing. Factors like low awareness about stool testing, discomfort with handling stool samples, challenges in sample transport and stigma were major barriers in its implementation.

**Conclusion:** Stool testing may help detect missed cases of TB among PW and U5C. It may be feasible to implement it, under the current National TB Elimination Programme (NTEP) framework, if there is adequate awareness among community and health care providers, and basic resources for its operationalization are met.

## Introduction

The World Health Organization (WHO) recommends that people with presumptive pulmonary TB, who can produce sputum, should be tested using molecular WHO recommended rapid diagnostic tests (mWRD) [1,2]. However, producing quality sputum samples may be challenging for many. In young children, the disease is often paucibacillary and spontaneous expectoration may not be feasible. Obtaining sputum samples from children may require procedures like gastric aspiration or induction of sputum, which is neither a pleasant experience for children nor an easy procedure to implement at low resource settings [3–5]. Pregnant Women (PW) may also face challenges in sputum production. During pregnancy, forceful coughing or clearing mucus may be distressing or may precipitate emesis, thereby preventing vigorous expectoration [6–8]. In such scenarios, stool testing for TB diagnosis can be a viable alternative. As respiratory secretions are often swallowed; *Mycobacterium tuberculosis (MTB DNA)* may survive passage through the gastrointestinal tract, and thus bacilli can be detected in stool [9,10]. In previous studies, molecular detection of *MTB* complex in stool samples using GeneXpert MTB/RIF has shown a diagnostic sensitivity of 60–70% compared to respiratory specimen cultures in both children and adults [11,12]. In Indian settings, especially the northern state of Bihar, which has constraints in context of TB care, infrastructure and trained manpower; the access to tertiary care services to diagnose is limited due to distance, travel cost and sparse distribution of diagnostic services, stool testing may play an important role in such settings [13–15]. Through this study, we aimed to assess whether stool testing increased microbiological confirmation of TB among PW and Under Five Children (U5C), and to explore factors influencing feasibility of its programmatic implementation in rural Bihar.

## Material & Methods

### Study Settings and Design

As per India TB report 2024, Bihar has one of the highest TB burdens in this Indian subcontinent, with an estimated incidence of 294 cases per 100,000 as compared to the national average of 195 cases 100,000 [16]. This was a community based sequential mixed-method study conducted between October 2023 and December 2024. The quantitative arm was used to estimate yield of Stool testing, followed by the qualitative arm, in which factors affecting implementation of Stool testing were explored. We implemented the study across five administrative blocks each from Begusarai (approximately 12,10,648 population) and Samastipur districts (approximately 15,65,810 population), of Bihar.

### Study Population

#### Quantitative

PW aged more than 18 years and U5C, residing in the intervention blocks receiving reproductive and childcare services at either Health & Wellness Centers (HWC) or Anganwadis, or through community-based home visits were symptomatically screened for TB. PW and U5C already on treatment for TB or those who completed treatment in the previous 6 months, admitted for other co-morbidities at the time of screening, not willing to consent for sputum or stool testing were excluded from the study. ***Qualitative*.**

For the qualitative study, PW, and guardians of the U5C who were diagnosed with TB by stool testing were included apart from the lab technician who tested the stool samples and Field Coordinators (FC) who worked as stool sample transporters.

### Sample Size

#### Quantitative

In collaboration with NTEP, Innovators In Health, India (IIH) supports active case finding of TB in the selected districts. All PW and U5C availing services at the different HWC-PHC/AAM and Anganwadi centers (childcare centers in India) in the intervention districts, during the study period were included as part of active case finding activities. As shown in figure 1, at the end of study period, among PW, 222 (60.2%) underwent sputum testing and 30 (8.1%) underwent stool testing while among under five children 279 (25.1%) underwent stool testing and sputum testing was done in 49 (4.4%) children

**Figure 1:**
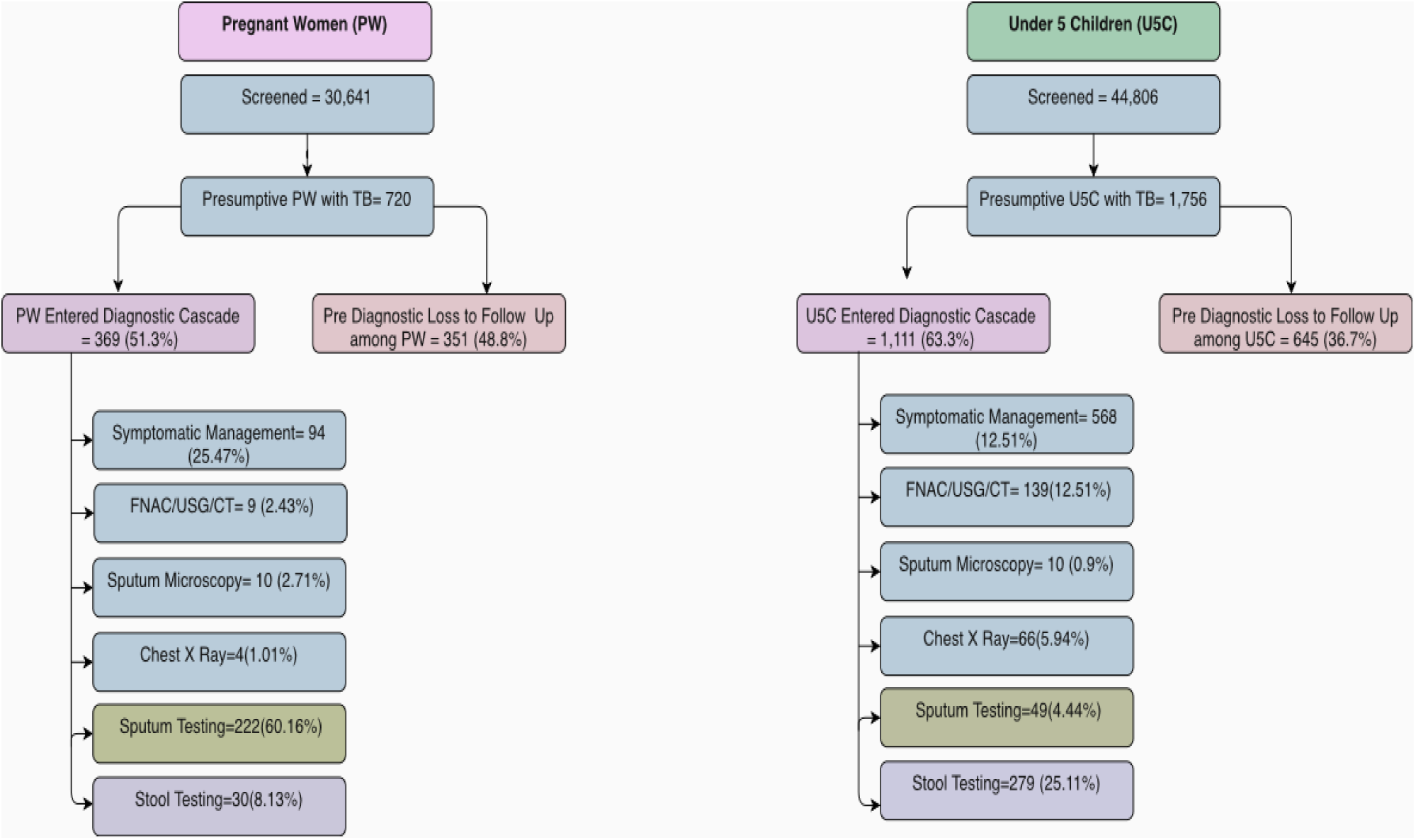
Total number of participants (PW & U5C) screened and tested. *FNAC= Fine Needle Aspiration Cytology, USG=Ultrasonography, CT=Computerized Tomography. Proportions of PW/U5C entering the diagnostic cascade/ Pre-Diagnostic Loss to Follow-Up, were estimated with Presumptives as denominator in each group. The individuals entering the diagnostic cascade were further stratified based on the management/diagnostic modality received*.

#### Qualitative

All the PW, diagnosed by stool testing and initiated on TB treatment, were approached, out of which two consented to participate in interviews. Similarly, among all the U5C, diagnosed by stool testing and initiated on treatment, five guardians of U5C consented for interview. Although 25-member teams of sample transporters were involved in stool collection, six of them were selected based on the maximum number of samples transported and consented for the interview. One laboratory technician, who was involved in testing of all the samples consented and was interviewed.

#### Data Collection Tools & methods Quantitative

PWs were mobilized by trained Accredited Social Health Activist (ASHA) workers to attend TB screening at reproductive and child health care clinics organized at the health facilities. For women, unable to visit these health facilities, home base screening was offered. For U5C TB screening was done at anganwadis and community settings. TB screening at respective facilities was performed and diagnostic linkage by field team members of IIH called as Field Coordinators (FC). The quantitative data was collected using a semi-structured, pre-tested and validated interview tool, containing socio demographic information, TB screening guide with questions on presence of i) cough more than 2 weeks, ii) sputum production, iii) hemoptysis iv) fever more than 2 weeks, v) night sweats, vi) chest pain, vii) significant weight loss, viii) presence of swollen glands in the body, and clinical follow-up details. Sputum samples were collected from each group and transported using cold chain for mWRD. For those unable to expectorate stool specimen were collected and tested. The detailed protocol for sample collection has been included as annexure 1 and lab testing procedure as annexure 2.

#### Qualitative

Qualitative data was collected using a semi-structured interview guide (Annexure 3), which was initially developed in English, then translated into Hindi and local Theta dialect. The tool was pre-tested among PwTB and FCs, before implementation in the field. For the PwTB (PW) and a guardian of the U5C, appointments were scheduled via phone after test results were delivered. The interviews for FCs and laboratory technicians were conducted during their routine working hours.

The interviews were conducted by three trained interviewers from the project team till theoretical saturation was reached. Each interview lasted 30-45 min long.

#### Data Analysis Quantitative

Data was entered into Microsoft Excel (Microsoft 365). After cleaning and management of missing data, a descriptive analysis was done using Jamovi (version 2.6.24). Testing modality-based yield was estimated, by dividing the total evaluated by specific diagnostic method (stool/sputum) by total number of bacteriologically confirmed PwTB.

#### Qualitative

Audio-recorded interviews were transcribed and translated into English by persons well-versed with the local language (Thethi in Bihar), Hindi and English languages. The data was analyzed manually using thematic analysis. After familiarization with the data, the initial coding was done by two of the investigators independently. The codes were grouped using axial coding to overarch global themes. This analysis was then reviewed by a third reviewer, to assess any bias in the interpretation of the findings. Any difference in opinion was resolved through discussion. Selected illustrative quotes were included as verbatims in this manuscript.

## Results

### Quantitative Results

Figure 2 shows that 252 PW and 328 U5C were evaluated for TB using sputum or stool as a sample on mWRD. Among PW, thirty stool samples and 222 sputum samples were evaluated. Stool testing yielded valid results in 96.7% samples, with MTB detected in 13.8% (4/29). Among U5C, 279 (85%) stool samples and 49 (15%) sputum samples were evaluated. Stool testing produced valid results in 96.1% samples, with MTB detected in 6.3% (17/268).

**Figure 2:**
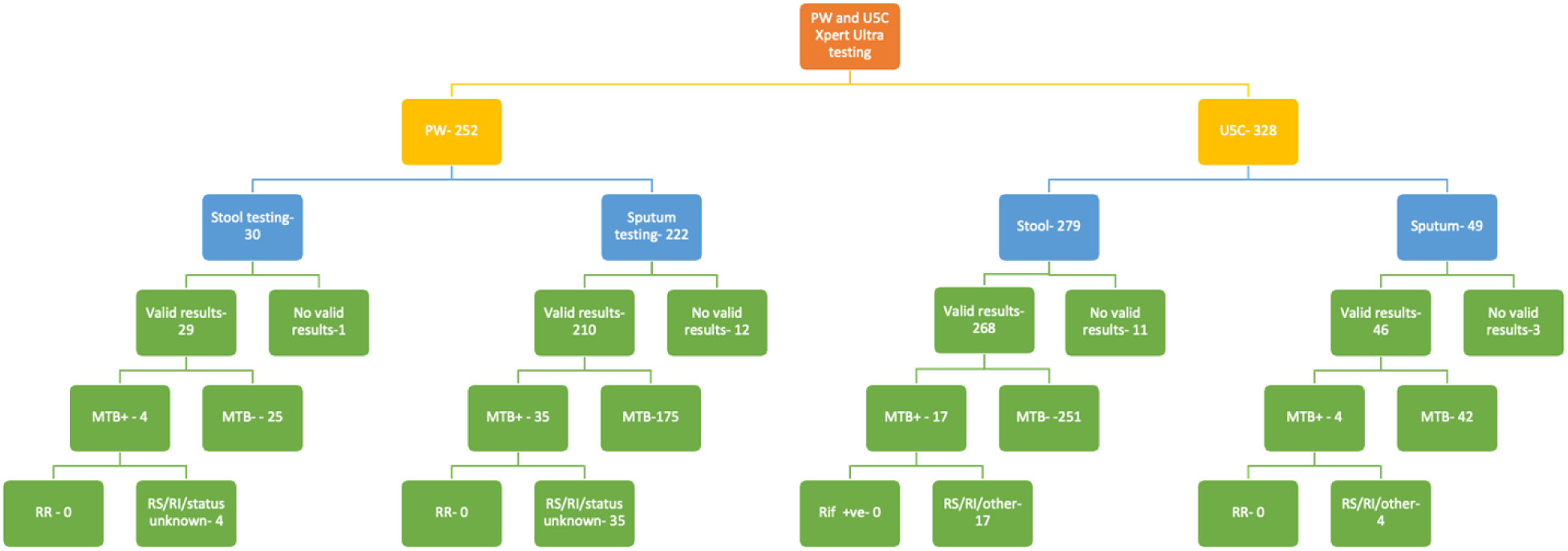
Flowchart describing the outcomes of sputum and stool testing in presumptive pregnant and under five children. PW-Pregnant Women, U5C-Under 5 Children, MTB: Mycobacterium Tuberculosis, RR: Rifampicin Resistance, RS: Rifampicin Sensitive, RI: Rifampicin Indeterminate

Among PW, stool testing contributed to an additional 4 MTB detections, equivalent to 10.3% (4/39) of all MTB positive results. Among U5C stool testing led to 17 MTB detection, corresponding to an additional yield of 80.9% (17/21). No rifampicin resistance (RR) was detected in both the groups.

### Qualitative Results

The interviews explored around the themes of i)Lack of awareness about stool testing for TB diagnosis, ii) *Discomfort, Disgust, and Fear Related to Stool Sample Collection iii)* Trust building in community through repeated personalized follow-ups iv) Stool Sample Quality and transport as a Challenge in Stool Testing v) Concerns on child’s health and pregnancy outcomes and the need for diagnosis and vi) Challenges after diagnosis with stool for TB and interaction with primary health care providers

### Lack of awareness about stool testing for TB diagnosis

The beneficiaries as well as the physicians, prescribing medications (as reported by PwTB), were unaware that stool can be used as a diagnostic sample for TB. Even though the report was being uploaded on an official government portal, they were not willing to accept the validity of the test. This led to initial resistance from PwTB as well as the physician ends of the care cascade.

***“We did not know about this stool test before. We only knew about the X-ray and sputum test.” (CG01)***

***“People were not aware about the stool test. They used to laugh at us.” (FC03)***

***“Initially doctors and the Nikshay portal have no idea about detection of TB through stool samples. They questioned the report. I opted to mention specimen as ‘other(s)’ for hustle free proceeding of treatment, just because we use ‘other(s),’ they didn’t question further” (LT)***

### Discomfort, Disgust, and Fear Related to Stool Sample Collection

While offering stool-based testing to presumptive PW and guardians of U5C, the IIH field team initially faced resistance, including ridicule and rejection. The FCs themselves expressed personal discomfort and hesitation regarding the handling and transportation of stool samples.

***“People used to laugh at me when they heard about the stool test. They told me that in the course of sputum collection, I started stool sample too.”(FC1)***

***“The mothers even complained that they would vomit even if they would try [to collect stool].” (FC6)***

***“The name’Stool’ sounds nice, as we were aware of its meaning. But when we came to know that it was a’latrine’ it made us feel uncomfortable.” (FC3)***

### Trust building in community through repeated personalized follow-ups

Families of the PW and U5C had never heard about stool testing for TB which is primarily a respiratory disorder. Hence it was essential to build trust and gain confidence, so that they provide the stool samples for testing’s gained the trust through multiple personal visits and engaging in discussion to clear their doubts.

***“Many a times we used to interact with them, isolating from the crowd as they felt uncomfortable in the crowd to discuss, first of all they were very hesitant for the stool collection as it was very uncommon to them. “After the proper counselling that the samples would go for diagnosis and proper medication would be carried out…then they turned-up. (FC4)***

***“He keeps on calling. He also visited to see how my son was doing. They suggested that I get the test done for my child.” (CG2)***

### Stool Sample Quality and Sample transport as a Challenge in Stool Testing

The quality of the stool samples determines the outcome of the tests, and this is the core concept ensuring the feasibility of incorporating stool in standard testing protocols. The lab technician highlighted that the challenges in testing were due to contamination, improper collection, and limited understanding of the caregivers on the sample collection. Insufficient samples led to invalid test results and repeat collection of stool samples. There were also challenges in collecting repeat samples. The team found it much easier to collect stool samples from PW than from U5C. The team had to reorganize their schedules, so that the sample reached the lab on time, without spillage or spoilage.

***“Undigested food particles in stool samples make samples non-diagnosable. We use the part of the stool sample where sputum-like consistency is visible. Too much odor indicates about the contaminated sample” (LT)***

***“Actually, there was no fixed time for the kids to pass their stools. “I had to make multiple visits for the sample collection because of this. In the process of collecting the sample from the field and then coming to the office took half of the day. It often became 6 pm to 7 pm while coming back from the lab.” (FC1)***

### Concerns on child’s health and pregnancy outcomes and the need for diagnosis

The caregivers expressed concerns about the ill health of their child and that treatment could not be started until a formal diagnosis for the disease was made. During the progression of the disease, and inability of the children to develop, they accepted the stool testing for TB diagnosis.

***“In case of kids, sputum does not get produced, so in this case stool test remains the only method*.**

***My child was very weak. I wanted to get my child cured.” (CG1)***

***“I was afraid about my pregnancy. I was worried about my baby; they told me medicines will be started later. (PW2)***

### Challenges after diagnosis with stool for TB and interaction with primary health care providers

PwTB diagnosed with stool samples, faced considerable stigma from primary health care providers, when they approached the public health system for initiation of treatment. There was lack of communication on the way forward, regarding the TB treatment cascade.

***“I was not briefed about the disease although I was given the report. They only told me that, I am positive. No, nobody explained to me about the disease. He did not even talk to me. The doctors behaved very rudely. They put off the fan and asked me to stand far from them.” (PW)***

## Discussion

Through this study we were able to demonstrate that stool testing can improve bacteriological confirmation of TB in PW with 10% and in U5C with more than 80%. We identified several factors that influenced the feasibility of its programmatic implementation in our setting such as lack of awareness of the test, people feeling uncomfortable collecting stools samples for testing and quality of the samples.

For both PW and U5C, we reported high rates of valid stool tests, 96.7 % and 96.1 %, respectively. The proportion of valid sputum tests was 94.5 % among PW and 93.8% among U5C. Although both the groups are not comparable in our study, higher valid stool tests may imply that for these groups, stools may be a better diagnostic sample than sputum requiring further exploration. Other studies conducted using stool Xpert have reported that high error/ non-valid test reports were a result of non-maintenance of cold chain, too less (<0.1g) or too much (> 0.8g) sample or delay in sample processing time [3,17]. Another issue highlighted was lack of standardized protocol and experience of the laboratory staff [3,18,19]. Initially, we also faced challenges in implementing stool testing in the field and keeping track of sample quality, transport, and monitoring. However, routine follow-up from the field team and adherence to protocols helped with high yield of the test results. Another factor, which may have contributed to higher yields, was use of the Petroff method for stool pre-processing, as compared to non-centrifugation-based methods like Simple One Step (SoS)/ Sucrose Flotation Methods used in other studies. These were achieved by a well-established laboratory infrastructure and trained lab technician [9,20,21].

Among PW, stool testing helped detect an additional 4 PwTB accounting for 10.3% of the total PwTB detected. These participants would have probably been missed out if sputum testing alone followed. This further strengthens the evidence that even for adult populations who find it unable/ difficult to expectorate, stool testing is an easy, non-invasive, and alternative diagnostic method [9,10,22,23]. Conventionally, stool testing has been explored for children, elderly and people living with HIV (PLHIV), our study adds the additional groups of PW.

Among U5C, stool testing detected an additional 17 PwTB among the total PwTB detected (21) in the group, which is a yield of 80.9%. This high yield further substantiates the utility of stool as a non-invasive diagnostic sample in children as recommended by WHO [9,10]. Studies conducted in many African countries, have repeatedly demonstrated, that stool testing increases the TB notification rates and enrollment of PwTB in TB diagnostic and treatment services specially for young children [9,24–26].

Qualitatively, we found that lack of awareness regarding stool testing for TB to be a major barrier for feasibility and uptake of the procedure. The awareness was low not only among beneficiaries, but also programmatic stakeholders as reported by the field and laboratory team. African countries, where stool testing was piloted like Mozambique and Nigeria, also reported low awareness levels, across the general population, PwTB, as well as health care providers [27–29]. Low awareness is a key barrier to acceptability as it can lead to higher rates of refusal. This can in turn lead to lower usage, which can prevent policy level stakeholders from enabling systems to train laboratory technicians and even prevent considering the status of the stool testing as valid.

We found that the field team found it challenging to accept the concept of stool collection and testing. Initially, they reported disgust, fear, and discomfort in handling the stool samples, even though they had received extensive training. Stool samples are often considered dirty, and this incites feelings of disgust. The malodorous nature of the stools can prevent checking the quality of samples, before transportation, and might lead to more erroneous results [27]. To ensure feasibility of the stool testing, a participatory approach is needed, to understand the perceptions of the sample transporters as well as the community. The field team also had more challenges collecting samples from U5C rather than adults, interfering in their routine household work and activities. Another study conducted in South Africa among older pediatric groups, also noted that children were reluctant and often embarrassed providing stool samples, which could be challenging if stool testing is expanded among children of older age groups [30]. These challenges are unique for U5C stool collection and transport protocols.

Accessible, confidential stool drop points or dedicated staff to increase the testing rates may be considered to tackle these challenges.

We also found that one of the reasons for acceptability of the stool testing was fear for the health of U5C and/or PW; as without diagnosis, treatment cannot be initiated. However, this perception is not sustainable for long term feasibility of any stool testing programme, only adequate levels of awareness, leading to informed acceptability can sustain utilization rates. Accessing stool testing for TB must be an active choice, rather than situational coercion.

We also found that, after being diagnosed with TB, irrespective of testing modality, PwTB faced significant stigma and discrimination from the health care providers. PwTB, after diagnosis, often goes through fear and emotional distrust and an additional burden of stigma from care providers, can lead to isolation and discriminatory practices, eventually affecting treatment outcomes [31–33]. Such individuals, who lose trust in primary care services, often hesitate to access care, appropriate linkage and in such instances maintenance of treatment becomes challenging. Because trust is important to introduce a new test and certainly a test which requires a stool sample. In our study, community trust building, through repeated counselling and follow-ups helped the sample collection. Showing sustained concerns motivates the beneficiaries to trust the test and have more satisfaction with testing procedure and results, which aids the prospects of acceptability and feasibility of the test [27,29].

While this study highlights the utility and factors affecting feasibility of stool tests, there are several limitations to this study. Sputum and stool groups were not compared parallelly, and the results were not validated against culture methods, so we cannot comment on the comparative yield of the two methods. Presumptive PW were not followed up, to assess pregnancy outcomes, which could reflect the impact of lead time gained in detection and treatment of TB. High pre-diagnostic loss to follow-up rates could have led to selection bias, among the participants entering the diagnostic cascade in this study. Pre-diagnostic loss to follow-up among both groups was high, which could have been due to various operational reasons like logistic related barriers, health system related barriers, and social factors. Linking them to diagnostic services would have helped us identify missed PwTB and further helped link more individuals to the TB care cascade.

All stool samples were tested using the GeneXpert device, whereas the most common testing modality available in the public health settings is Truenat by Molbio. Further studies are needed to assess the feasibility of stool testing using routinely available Truenat devices. While the Petroff method was used in this study for stool pre-processing, it utilizes reagents and centrifuge for processing of samples. Unlike WHO recommendation of non-centrifugation methods, this protocol could lead to additional resource burden and costs if replicated in the current public laboratory infrastructure.

One of the limitations of the qualitative aspect is that the study does not explore the opinion of physicians initiating PwTB on treatment.

## Conclusion

Our study demonstrated that stool-based testing for TB contributes to a higher proportion of bacteriologically confirmed TB among U5C and PW, although in the later group the increase was much lower. While in the study it was feasible to conduct stool testing for detection of TB, several factors may limit its feasibility, when scaled up.

## Data Availability Statement

The datasets used and/or analyzed during the current study are available from the corresponding author on reasonable request.

## Ethical Consideration

The study was approved by the Institutional Ethics Committee (IEC) of Indira Gandhi Institute of Medical Sciences (IGIMS), Patna vide letter no. 1209/IEC/IGIMS/2023. Data was collected from the participants after administration of the patient information sheet and obtaining informed consent. Parental consent was obtained for all U5C. All participants were allotted a unique ID, and the data was de-identified prior to analysis for anonymization and maintaining confidentiality.

## Author Contributions

Kajal Taluja and Shailja Shah contributed to conceptualization and methodology. Manish Kumar, Manish Bharadwaj and Miranda Brouwer provided supervision at all stages of the study. Kajal Taluja, Shailja Shah, Arup Saha, Chandan Mahadevan and Navnit Nisha contributed to data curation, formal analysis, and writing of the original draft. Tushar Garg, Rakesh Kumar, Sweta Muni, contributed to validation, interpretation of the findings, and writing – review and editing.

Kajal Taluja, Shailja Shah, Mithilesh Kumar, Ranjeet Kumar Sharma, Onkar Nath, Navnit Nisha, Nazreen Nazir, Chandan Mahadevan, Bal Krishna Mishra, and Rakesh Kumar contributed to investigation, project administration, resources, and data acquisition during field operations. Manish Kumar and Manish Bhardwaj contributed to overall project administration.

All authors contributed to writing – review and editing, approved the final version of the manuscript, and agreed to be accountable for all aspects of the work.

## Funding Statement

This work was supported by the Stop TB Partnership/ TB Reach Grant Wave 10. (STBP/TBREACH/GSA/W10-10266) However, it had no role in the conceptualization, design, data collection, analysis, decision to publish, or preparation of the manuscript.

## Competing Interests

Tushar Garg is employed by Stop TB Partnership and has been involved in review of the manuscript. The other authors declare no conflict of interest.

## Acknowledgements

The authors extend their sincere gratitude to the National Tuberculosis Elimination Programme (NTEP) Bihar for their invaluable support of this research. They also deeply appreciate the participation and cooperation of the pregnant women, guardians of children, technician and field team who participated in the interview. Furthermore, authors express their thanks to the Indira Gandhi Institute of Medical Sciences (IGIMS) for granting ethical board approval for this study, and to its core donor, the STOP TB Partnership and TB Reach, for their crucial support and encouragement in this innovative endeavor. The authors also appreciate the technical support received by the Communicable Disease Officer of Samastipur and Begusarai districts. Finally, the authors acknowledge the contributions of the entire project team and WHO consultants Dr. Greevarn Raikwar and Dr. Umar Aquil.

## Notes

### Author Declarations

The study was approved by the Institutional Ethics Committee (IEC) of Indira Gandhi Institute of Medical Sciences (IGIMS), Patna vide letter no. 1209/IEC/IGIMS/2023.

